# COVID-19 preventive behaviors among people with anxiety and depression: Findings from Japan

**DOI:** 10.1101/2020.06.19.20135293

**Authors:** Andrew Stickley, Tetsuya Matsubayashi, Hajime Sueki, Michiko Ueda

## Abstract

Little is known about COVID-19 preventive behaviors among individuals with mental health problems. This study used cross-sectional online survey data from 2000 Japanese adults collected in April and May, 2020, to examine the association between anxiety and depression and COVID-19 preventive behaviors. Results from logistic regression analyses showed that both anxiety and depression were associated with lower odds for engaging in preventive behaviors such as wearing a face mask and hand washing. Our results highlight the importance of facilitating the performance of preventive behaviors in individuals with mental health problems to prevent the spread of COVID-19 in this population.

## 1. Introduction

It has been suggested that individuals with serious mental illness may be especially at risk for coronavirus exposure and infection (Shinn and Viron, 2020). In particular, the effects of factors such as socioeconomic disadvantage and lower health literacy, as manifest in a lack of understanding of preventive health behaviors (Kim et al., 2019; Shinn and Viron, 2020), may be compounded by specific characteristics associated with mental disorders, such as hopelessness, that may hinder adherence to necessary health behaviors (Gehi et al., 2005). Difficulty in undertaking COVID-19-related preventive behaviors among those with mental health problems may have been further exacerbated during the ongoing pandemic by the novel nature of the disease itself, and the plethora of information (‘infodemic’) it has generated, both in conventional and social media, that has resulted in uncertainty and worry (Fiorillo and Gorwood, 2020).

Against this backdrop, the current study will examine the effects of anxiety and depression on COVID-19 preventive behaviors in a sample of the Japanese general population. As yet, there has been little research on the association between mental health and infectious disease-preventive behaviors and the research that has been undertaken has produced mixed results. An earlier study from Hong Kong found that compared to those with low anxiety, individuals with high, and especially moderate levels of anxiety had significantly higher odds for adopting ≥ 5 precautionary measures against severe acute respiratory syndrome (SARS) (Leung et al., 2003). In contrast, a recent study from China reported that anxiety was not related to any differences in the adoption of preventive measures while depression was associated with fewer preventive measures being taken in response to the COVID-19 pandemic (Liu et al., 2020).

A focus on Japan may be particularly instructive. Even though the impact of the COVID-19 pandemic has been less severe than in many countries – at least in terms of mortality – some research has indicated that its effects on wellbeing may have been comparatively greater than in other countries. For example, a recent international poll showed that 86% of the Japanese population was afraid of catching the virus; this was a higher percentage than in other high-income countries such as the United States and United Kingdom where approximately 60% of the population reported that they were scared of contracting COVID-19 (YouGov, 2020). Importantly, a recent study has reported that although the vast majority of Japanese adults have adopted preventive measures, around 20% of the working-age population (age 20-64) remain reluctant to do so (Muto et al., 2020). However, that study did not focus specifically on the possible effects of mental health in the adoption/non-adoption of preventive behaviors.

## 2. Methods

### 2.1. Participants

Two rounds of an online survey of the Japanese population were administered between April 16 and April 18, 2020 (1^st^ round) and May 15 and May 17 (2^nd^ round). A commercial survey company, the Survey Research Center, was tasked to send out a set of screening questions to approximately 10,000 respondents from its commercial web panel and then to construct a sample of 1,000 respondents based on their demographic characteristics in each round. A new set of respondents was drawn in the second round. The final sample comprised respondents who were representative of the Japanese general population in terms of the area of their residency, sex, and age distribution. The respondents in the final sample answered online questions about their mental health, personal economic situation, and preventive behavior regarding COVID-19, among others. The final sample size was 2,000.

### 2.2. Ethics statement

This study was approved by the Ethics Committee of Waseda University (approval case number: 2020-050) and Osaka School of International Public Policy, Osaka University. The survey participants were informed of the purpose of the study prior to their participation and had the option to quit the survey at any time. The respondents provided explicit consent that the information they provided could be used for the purpose of this study. The data are completely anonymous.

### 2.3. Mental health status

The self-report Patient Health Questionnaire (PHQ-9) was used to assess past-week depression (Spitzer et al., 1999). A score of 10 or above (out of 27) was regarded as a case of at least moderate depressive symptomatology (Levis et al., 2019). The Cronbach’s alpha value for the scale was 0.90. The self-administered 7-item Generalized Anxiety Disorder Scale (GAD-7) was used to measure past two-week anxiety (Spitzer et al., 2006). A score of 10 or higher (out of 21) was regarded as a case of at least moderate anxiety disorder (Lowe et al., 2008). The scale had a good degree of internal reliability (Cronbach’s alpha = 0.92).

### 2.4. COVID-19 preventive health behaviors

Information was collected on 13 COVID-19 preventive behaviors (no/yes). The specific behaviors and their prevalence are detailed in Table 1.

**Table 1.**
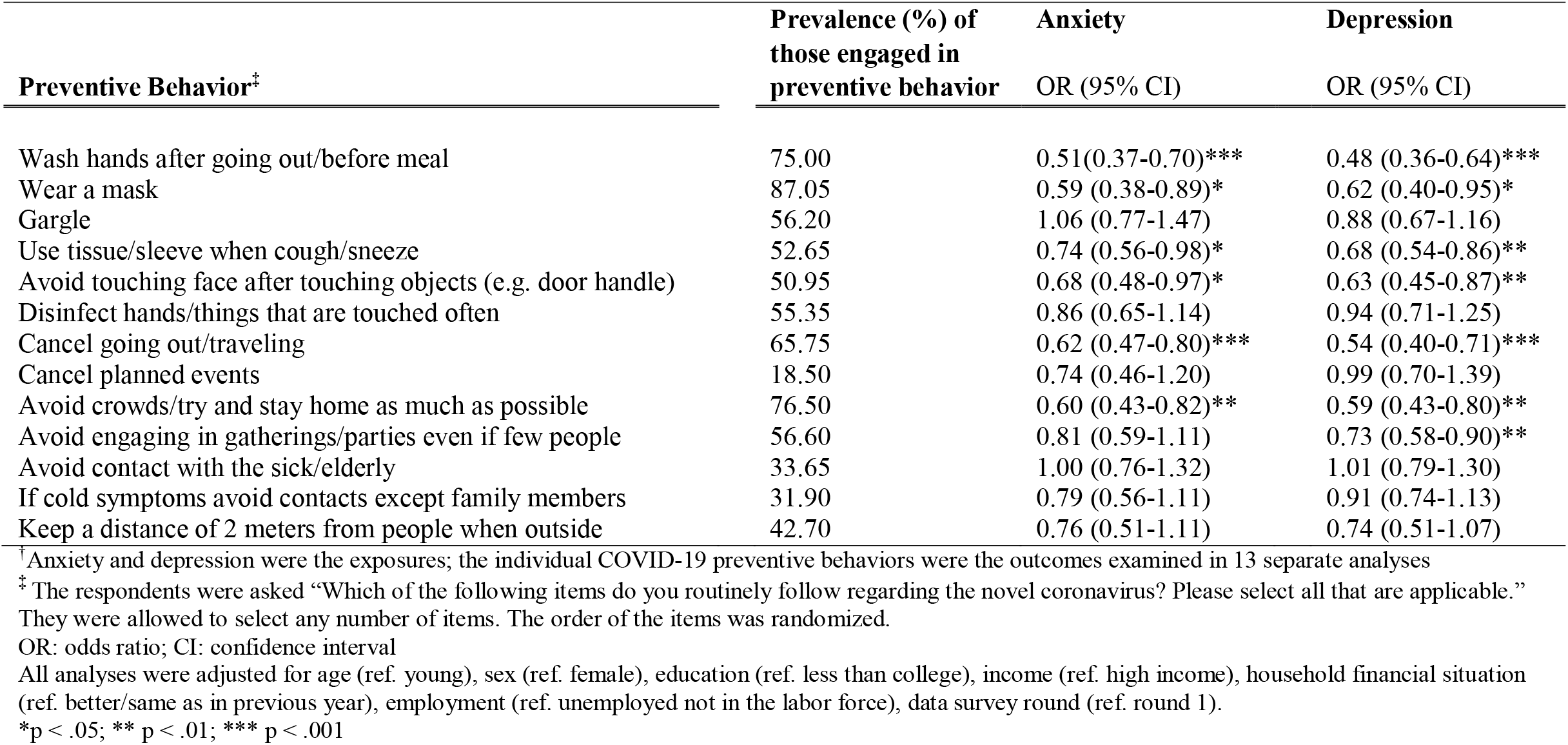
Association between anxiety and depression and individual COVID-19 preventive behaviors among Japanese adults^†^

| Preventive Behavior<sup>‡</sup> | Prevalence (%) of<br>those engaged in<br>preventive behavior | Anxiety<br>OR (95% CI) | Depression<br>OR (95% CI) |
| --- | --- | --- | --- |
| Wash hands after going out/before meal | 75.00 | 0.51(0.37-0.70)*** | 0.48 (0.36-0.64)*** |
| Wear a mask | 87.05 | 0.59 (0.38-0.89)* | 0.62 (0.40-0.95)* |
| Gargle | 56.20 | 1.06 (0.77-1.47) | 0.88 (0.67-1.16) |
| Use tissue/sleeve when cough/sneeze | 52.65 | 0.74 (0.56-0.98)* | 0.68 (0.54-0.86)** |
| Avoid touching face after touching objects (e.g. door handle) | 50.95 | 0.68 (0.48-0.97)* | 0.63 (0.45-0.87)** |
| Disinfect hands/things that are touched often | 55.35 | 0.86 (0.65-1.14) | 0.94 (0.71-1.25) |
| Cancel going out/traveling | 65.75 | 0.62 (0.47-0.80)*** | 0.54 (0.40-0.71)*** |
| Cancel planned events | 18.50 | 0.74 (0.46-1.20) | 0.99 (0.70-1.39) |
| Avoid crowds/try and stay home as much as possible | 76.50 | 0.60 (0.43-0.82)** | 0.59 (0.43-0.80)** |
| Avoid engaging in gatherings/parties even if few people | 56.60 | 0.81 (0.59-1.11) | 0.73 (0.58-0.90)** |
| Avoid contact with the sick/elderly | 33.65 | 1.00 (0.76-1.32) | 1.01 (0.79-1.30) |
| If cold symptoms avoid contacts except family members | 31.90 | 0.79 (0.56-1.11) | 0.91 (0.74-1.13) |
| Keep a distance of 2 meters from people when outside | 42.70 | 0.76 (0.51-1.11) | 0.74 (0.51-1.07) |

<sup>†</sup>Anxiety and depression were the exposures; the individual COVID-19 preventive behaviors were the outcomes examined in 13 separate analyses<sup>‡</sup>The respondents were asked “Which of the following items do you routinely follow regarding the novel coronavirus? Please select all that are applicable.” They were allowed to select any number of items. The order of the items was randomized.
OR: odds ratio; CI: confidence interval
All analyses were adjusted for age (ref. young), sex (ref. female), education (ref. less than college), income (ref. high income), household financial situation (ref. better/same as in previous year), employment (ref. unemployed not in the labor force), data survey round (ref. round 1).
\*p < .05; \*\* p < .01; \*\*\* p < .001

### 2.5. Covariates

Information was also obtained on age, sex, education, income, household financial situation (versus previous year), employment status and data survey round.

### 2.6. Statistical analysis

Logistic regression analysis was performed to examine the association between anxiety and depression and preventive behaviors. Two analyses were performed. In the first analysis the association between anxiety and depression and each of the individual preventive behaviors was examined using binomial logistic regression. In the second analysis a combined preventive behavior score variable was created by summing each preventive behavior score and ordered logistic regression analysis was used to examine the associations. All analyses were adjusted for the above-listed covariates. The standard errors were heteroskedasticity-robust, and clustered by prefecture. The analysis was conducted using STATA/MP (version 16, Stata Corporation, College Station, TX). The results are presented as odds ratios (OR) with 95% confidence intervals (CI). The level of statistical significance was set at p < 0.05 (two-tailed).

## 3. Results

The prevalence of anxiety was 10.9%, while 17.3% of the respondents had depressive symptoms. For anxiety, ORs were negative for 11 of the 13 preventive behaviors (Table 1). Individuals with anxiety were significantly less likely to engage in six of the preventive behaviors. Specifically, they had a 40-49% reduction in odds for washing hands, wearing a mask and avoiding crowds, and a 26-38% reduction in odds for using a tissue/sleeve when coughing/sneezing, avoid touching face and cancel going out. Depression was also associated with significantly reduced odds for the same six preventive behaviors. In addition, depression was also associated with a 27% reduction in the odds for avoiding engaging in gatherings (OR: 0.73, 95%CI: 0.58-0.90). Both anxiety (OR: 0.65, 95%CI: 0.48-0.88) and depression (OR: 0.64, 95%CI: 0.47-0.87) were associated with significantly reduced odds for engaging in all of the preventive behaviors combined (Table 2).

**Table 2.** Association between anxiety and depressive symptoms and all COVID-19 preventive behaviors combined among Japanese adults^†^

|  | **Anxiety** | **Depression** |
| --- | --- | --- |
|  | **OR (95%CI)** | **OR (95%CI)** |
| Preventive Behaviors | 0.65 (0.48-0.88)** | 0.64 (0.47-0.87)** |

<sup>†</sup>Anxiety and depressive symptoms were the exposures; COVID-19 preventive health behaviors were the outcomes OR: odds ratio; CI: confidence interval
Both analyses were adjusted for age (ref. young), sex (ref. female), education (ref. less than college), income (ref. high income), household financial situation (ref. better/same as in previous year), employment (ref. unemployed not in the labor force), data survey round (ref. round 1)
\*\* p < .01

## 4. Discussion

This study used data from 2000 Japanese adults collected in April and May 2020 to examine the association between mental health status and COVID-19 preventive behaviors. Results showed that individuals with anxiety and depression were significantly less likely to engage in a variety of preventive behaviors including hand washing, wearing a mask, avoiding crowds/staying at home and taking measures to control the effects of coughing/sneezing.

Until now there has been little focus on the association between mental health and protective behaviors pertaining to the prevention of infectious disease although our results agree in part with those from a recent study from China, which showed that depressive symptoms may inhibit preventive behaviors in response to the COVID-19 pandemic (Liu et al., 2020). It is uncertain what underlies lower odds for engaging in preventive behaviors. A recent review article has highlighted the possible role of disorganized thinking in individuals with mental illness that may prevent a full understanding of the gravity of the current situation and measures to prevent the spread of the virus (Hamada and Fan, 2020).

Importantly, these results indicate that people with mental health problems may be at increased risk for COVID-19 infection given their lower engagement in a number of preventive behaviors. This highlights the importance of maintaining and facilitating mental health consultation services during the ongoing pandemic – irrespective of the increasing difficulties associated with lockdown/travel restrictions (Yao et al., 2020). They also emphasize the importance of educating patients about the dangers of COVID-19 and how to protect themselves against the virus (Shinn and Viron, 2020). Finally, our findings also suggest that further research on the effects of COVID-19 among individuals with mental health problems is now urgently warranted.

## Data Availability

The data used in this study are not available due to the sensitive nature of the data.

## Notes

### Competing Interest Statement

The authors have declared no competing interest.

### Funding Statement

This work was financially supported by JSPS Grants-in-Aid for Scientific Research Grant Number 20H01584. The funders had no role in study design, data collection and analysis, decision to publish, or preparation of the manuscript.

### Author Declarations

This study was approved by the Ethics Committee of Waseda University (approval case number: 2020-050) and Osaka School of International Public Policy, Osaka University.

